# Analytical and Clinical Performance of the Panbio COVID-19 Antigen-Detecting Rapid Diagnostic Test

**DOI:** 10.1101/2020.10.30.20223198

**Authors:** Andrea Alemany, Bàrbara Baro, Dan Ouchi, Maria Ubals, Marc Corbacho-Monné, Júlia Vergara-Alert, Jordi Rodon, Joaquim Segalés, Cristina Esteban, Gema Fernandez, Lidia Ruiz, Quique Bassat, Bonaventura Clotet, Jordi Ara, Martí Vall-Mayans, Camila G-Beiras, Ignacio Blanco, Oriol Mitjà

## Abstract

**Background:** The current standard for COVID-19 diagnosis, RT-qPCR, has important drawbacks for its use as a tool for epidemiological control, including the need of laboratory-processing, high cost, and long turnaround from sampling to results release. Antigen-based rapid diagnostic tests (Ag-RDT) provide a promising alternative for this purpose.

**Methods:** We assessed the analytical and clinical performance of the Ag-RDT Panbio COVID-19 Ag Test (Abbott), using RT-qPCR as a reference test. The clinical performance was assessed using nasopharyngeal swabs, collected in routine practice for case confirmation and contact tracing, and nasal mid-turbinate swabs, collected in preventive screenings of asymptomatic individuals. Fresh samples were analysed by RT-q-PCR, stored at -80 °C, and analysed using the Ag-RDT according to the manufacturer instructions.

**Findings:** The Ag-RDT had a limit of detection of 6·5×10^5^ copies/reaction. The clinical performance was assessed on 1,406 frozen swabs with a PCR result available: 951 (67·7%) positive and 455 (32·4%) negative. The Ag-RDT identified the presence of SARS-CoV-2 in 872 of 951 PCR-positive samples (91·7%; 95% CI 89·8-93·4 and ruled out its presence in 450 of 455 PCR-negative samples (specificity 98·9%; 95% CI 97·5– 99·6). Sensitivity increased in samples with lower Ct values (Ct <25, 98·2%; Ct<30, 94·9%) and was higher among symptomatic cases (92·6%) and their contacts (94·2%) than among asymptomatic individuals (79·5%). In the setting of asymptomatic screening, sensitivity also increased with lower Ct values (Ct <25, 100%; Ct<30, 98·6%). Assuming a pre-test probability of 5%, the negative and positive predictive values were 99·6% (99·5 – 99·6) and 81·5% (65·0 – 93·2), respectively.

**Interpretation:** The Panbio COVID-19 Ag-RDT has high sensitivity for detecting the presence of SARS-CoV-2 in nasal or nasopharyngeal swabs of both, symptomatic and asymptomatic individuals. The diagnostic performance of the test is particularly good in samples with viral loads associated with high risk of viral transmission (Ct <25), which show high positive and negative predictive values even when assuming a prevalence as low as 5%.

**Funding:** Blueberry diagnostics, Fundació Institut d’Investigació en Ciències de la Salut Germans Trias i Pujol, and #YoMeCorono.org crowfunding campaing.

**Research in context:** *Evidence before this study:* On October 6, 2020, we searched PubMed for articles containing “Antigen”, “test”, “SARS-CoV-2”, “COVID-19” and “performance” in either the title or the abstract. We found five studies that showed the accuracy of point-of-care tests in identifying SARS-CoV-2 antigens for confirmation of clinically suspected COVID-19. We found high variability in the diagnostic accuracy of Ag-RDT. Most tests showed high specificity (i.e., 99% or higher), whereas sensitivity ranged from 11% to 92%; only one test reported sensitivity higher than 60%. We found no studies investigating the diagnostic accuracy of the *Panbio COVID-19 Ag Test*. We found no studies that assessed the performance of Ag-RDT for population-level screening of asymptomatic individuals.

*Added value of this study:* Our analysis provides information regarding the diagnostic accuracy of the *Panbio COVID-19 Ag Test* when tested on 1,406 frozen samples of nasopharyngeal and nasal swabs collected in routine practice for diagnostic confirmation of symptomatic individuals with suspected COVID-19 or contacts exposed to a positive case, and preventive screenings of unexposed asymptomatic individuals. Compared with RT-qPCR as reference test, the Ag-RDT showed a sensitivity and specificity of 91·7% and 98·9%. Test sensitivity increased in samples with viral load associated with high risk of transmission (Ct <25), reaching more than 98%, regardless of the presence of symptoms.

*Implications of all the available evidence:* Available evidence show variability in the diagnostic performance of marketed Ag-RDT. Our results provide substantial evidence that the point-of-care *Panbio COVID-19 Ag Test* can accurately identify SARS-CoV-2 antigens in people with suspected clinical COVID-19 as well as in asymptomatic people with high viral load and therefore, associated with higher risk of transmission. This finding represents a potentially useful advance for mass screening of asymptomatic people at the point-of-care.

## Introduction

Strategies for early identification of severe acute respiratory syndrome coronavirus 2 (SARS-CoV-2) cases and their contacts are a mainstay for containing the viral spread. Reverse transcription-polymerase chain reaction (RT-qPCR) analysis of nasopharyngeal swabs remains the gold standard for identifying the presence of the SARS-CoV-2 genome. However, the need for laboratory-processing, high cost, and long turnaround from sampling to results release limits the feasibility of this technique for community-based testing strategies. The growing body of evidence on the lack of infectivity of cases with low viral load^1,2^ suggests that frequent testing with rapid diagnostic tests Antigen-detecting RDTs (Ag-RDT)-even those with low sensitivity-may be more adequate than RT-qPCR for epidemiological control of the SARS-CoV-2.^3^ Furthermore, preliminary data on the performance of Ag-RDTs suggest that their sensitivity increases with the viral load, reaching 99% when testing specimens with high viral loads.^4,5^

Ag-RDTs, commonly used in the diagnosis of infectious respiratory diseases, have recently become available for the identification of SARS-CoV-2.^6^ Tests designed for clinical diagnosis of symptomatic people require high analytic sensitivity. In contrast, test requirements^7^ for a containment strategy of COVID-19 are rapid turnaround time (i.e., less than 20 minutes), low cost, and ease-of-use to allow frequent testing at the point-of-care.^8–12^

Currently, the WHO recommends using Ag-RDTs to support diagnosis of cases and contacts during outbreak investigations and monitor trends in disease incidence, particularly in remote settings or closed groups (e.g., schools, care homes, or prisons), but not to screen asymptomatic populations.^6^ In this study, we investigated the performance of a SARS-CoV-2 Ag-RDT for confirmation of clinically suspected COVID-19 and their contacts using nasopharyngeal specimens, and for community screening of asymptomatic people using nasal mid-turbinate specimens.

## Methods

### Test selection

After a literature review and web search^13^ for SARS-CoV-2 rapid antigen-based tests, we selected four candidates deemed potentially suitable for diagnosis and screening strategies: COVID-19 Ag Respi-Strip (Coris BioConcept, Gembloux, Belgium), Standard Q COVID-19 Ag Test (SD Biosensor, Suwon, South Korea), Standard F COVID-19 Ag FIA (SD Biosensor, Suwon, South Korea), and Panbio^™^ COVID-19 Ag Test (Abbott Laboratories, Illinois, USA). The selected tests were pre-screened by triplicate on 40 frozen samples of nasopharyngeal swabs with known PCR result (Table S1, Supplementary Material). Based on the pre-screening results, the Panbio COVID-19 Ag Test was selected for the assessment of analytical and clinical performance.

The Panbio™ COVID-19 Ag Rapid Test Device is a chromatographic, immunoassay-based platform. For a positive result, a gold conjugate human IgG specific to SARS-CoV-2 Ag and anti-SARS-CoV-2 antibody form a test line in the result window.

### Analytical performance of antigen rapid tests

The analytical performance of the Ag-RDT test was assessed using a SARS-CoV-2 isolate (ID EPI_ISL_510689) propagated in Vero E6 cells (ATCC CRL-1586). The median tissue culture infective dose (TCID_50_), defined as the dilution that caused cytopathic effect in 50% of the inoculated cell cultures, was calculated by titrating a passage 3 SARS-CoV-2 stock into Vero E6 cells and culturing them at 37 °C in a 5% CO_2_ incubator for six days. Cells were cultured in Dulbecco’s modified Eagle medium (DMEM; Lonza) supplemented with 5% foetal calf serum (FCS; EuroClone), 100 U/mL penicillin, 100 µg/mL streptomycin, and 2 mM glutamine (all ThermoFisher Scientific).

The analytical sensitivity was estimated by testing in triplicate eight ten-fold serial dilutions of the SARS-CoV-2 suspension in PBS (Lonza). All samples were cross-validated by RT-qPCR and virus isolation in Vero E6 cells. RT-qPCR for genomic detection (UpE assay; Corman et al., 2020) was conducted from viral RNA extracted using the Indimag Pathogen kit (Indical Biosciences) from all serial dilutions on a Biosprint 96 workstation (Qiagen). A plasmid containing the complete envelope gene (GenBank NC_045512.2; IDT, Inc.) was used to quantify the amount of SARS-CoV-2 genome copies of each sample. Virus isolation was performed in 16 replicates by inoculating 50 µL of each replicate per well (containing15,000 Vero E6 cells, seeded in 96-well plates), and incubating for 1 h at 37°C in a 5% CO_2_ atmosphere. Then, 100 µL of supplemented DMEM were added to each well and the plates were maintained at 37°C and 5% CO_2_. Plates were daily monitored under the light microscope and wells were evaluated for the presence of cytopathic effect for 6 days.

### Clinical performance

The clinical performance of the Ag-RDT was assessed using RT-qPCR as a reference test with a cycle threshold (Ct) < 40 as the criteria for a positive result. Samples consisted of nasopharyngeal swabs collected in routine practice for diagnostic confirmation of symptomatic individuals with suspected COVID-19 or contacts exposed to a positive case, and nasal mid-turbinate swabs collected in preventive screenings of unexposed asymptomatic individuals in the general population. Swab specimens were placed into sterile tubes containing viral transport media (DeltaSwab Virus, Deltalab, Ref 304301). RT-qPCR tests were performed on fresh samples stored at 2 – 8 °C for up to 72 hours; samples were then stored at –80 °C until their use for Ag-RDT.

Samples were classified according to the disease status of the participant (i.e., suspected symptomatic case, exposed asymptomatic contact, and unexposed asymptomatic individual) based on the national guideline definitions. Individuals’ data were collected anonymously and handled according to the General Data Protection Regulation 2016/679 on data protection and privacy for all individuals within the European Union and the local regulatory framework regarding data protection.

RNA for RT-qPCR tests were extracted from fresh samples using the viral RNA/Pathogen Nucleic Acid Isolation kit, optimized for a KingFisher instrument according to the manufacturer’s instructions. PCR amplification was conducted according to the recommendations of the 2019-nCoV RT-qPCR Diagnostic Panel of the Centers for Disease Control and Prevention (CDC).^14^ Briefly, a 20 μL PCR reaction mix was prepared that contained 5 μL of RNA, 1·5 μL of N3 primers and probe (2019-nCov CDC EUA Kit, catalog no. 10006770, Integrated DNA Technologies), and 5 μL of TaqPath 1-StepRT-qPCR Master Mix (Thermo Fischer). Thermal cycling was performed on either Applied Biosystems 7500 or QuantStudio5 Real-Time PCR instruments (Thermo Fischer) at the following conditions: 15 min at 50 °C for reverse transcription, followed by 2 min at 95°C, and then 45 cycles of 3 sec at 95°C and 30 sec at 55°C.

Rapid antigen tests were performed according to the manufacturer’s IFU (Abbott, Illinois, USA) except for the use of a viral transport media (DeltaSwab Virus) and swab storage as a frozen specimen. Internal validation showed no significant change in the test performance using Abbot test Kit buffer or a mix of the Kit buffer and transport media at 1:3 dilution; likewise, the use of frozen specimens showed no significant differences compared with fresh ones. All Ag-RDT determinations were performed by two blinded technicians, who used 100 μL of 1:3 mix of the Kit buffer and the sample previously thawed and homogenized. Samples were applied directly to the test cassette and incubated for 15 minutes at room temperature before reading results at naked eye, according to the manufacturer instructions (i.e., the presence of any test line (T), no matter how faint, indicates a positive result).

### Statistical Analysis

We determined the sample size needed to estimate sensitivity with 80% power and precision 2·25% was 944 if the actual sensitivity of the index test was 93·5% (reported by the manufacturer) and specificity with 80% power and 2·25% precision was 450 if the actual specificity was 99·6% (reported by the manufacturer).

Sensitivity and specificity were estimated as defined by Altman et al.^15^, and reported as a percentage and the exact binomial 95% confidence interval (CI). The negative-predictive value (NPV) and positive-predictive value (PPV) were estimated by considering the prevalence as pre-test probability;^16^ the two values were modelled for pre-test probabilities ranging between 0·05 and 0·9 and plotted with the exact binomial 95% CI.^17^ The significance threshold was set at a two-sided alpha value of 0·05. All analyses and plots were performed using R version 3·6^18^.

## Results

The initial viral stock of SARS-CoV-2 used for analytical performance was titrated on Vero E6 cells, obtaining a 10^6·4^ TCID_50_/mL. This value corresponded to an average viral load of 6·8 × 10^8^ genome copies/reaction measured by RT-qPCR. The Ag-RDT yielded positive results in viral stock dilutions of 1:10^3^ and higher, corresponding to a limit of detection (LoD) of 6·5 × 10^5^ genome copies/reaction as assessed by RT-qPCR (Table 1).

**Table 1.**
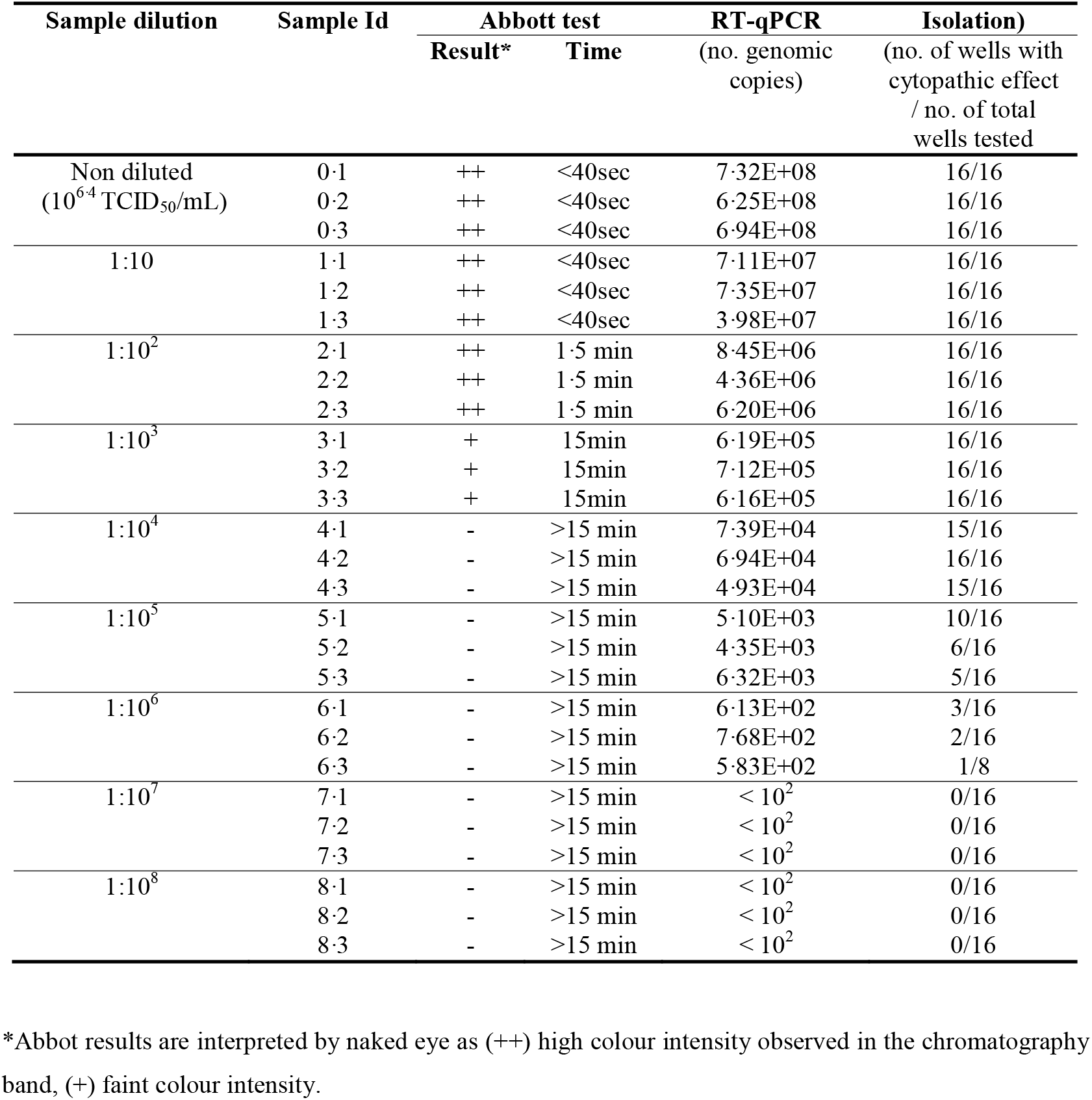
Results of the analytical performance assessment

The clinical performance was analysed on 1,406 frozen swabs with a RT-qPCR result available: 951 (67·6%) positive and 455 (32·4%) negative. The mean age of the sampled individuals was 40·4 years, and 936 (66·6%) were female (Table 2). The Ct value was <20 in 258 (18·3%) samples, 20-to-24 in 305 (21·7%), 25-to-29 in 285 (20·3%), and >30 in 103 (7·3%). Nasopharyngeal swabs had been collected in the setting of diagnosis confirmation in symptomatic cases (446/1406, 31·7%) and contact tracing (473/1406, 33·6%), and mid-turbinate nasal swabs were collected in mass screening campaigns (487/1406, 34·6%).

**Table 2.** Baseline characteristics of sampled individuals.

|  | **All samples** | **PCR+** | **PCR-** |
| --- | --- | --- | --- |
| **N** | 1406 | 951 | 455 |
| **Age** (years), *mean (SD)* | 40.40 (24.50) | 43.05 (24.88) | 34.90 (22.75) |
| **Gender**, *n (%)* (n=1,389) |  |  |  |
| *Male* | 453 (32.2) | 261 (27.9) | 192 (42.4) |
| *Female* | 936 (66.6) | 675 (72.1) | 261 (57.6) |
| **Ct value**, *median (IQR)* (n=951) | 23.63 [19.72, 27.31] | 23.63 [19.72, 27.31] | .. |
| **Ct stratification**, *n (%)* (n=951) |  |  |  |
| <20 | 258 (18.3) | 258 (27.1) | .. |
| 20-24 | 305 (21.7) | 305 (32.1) | .. |
| 25-29 | 285 (20.3) | 285 (30.0) | .. |
| >30 | 103 (7.3) | 103 (10.8) | .. |
| **Disease status**, *n (%)* |  |  |  |
| *Case* | 446 (31.7) | 419 (44.1) | 27 (5.9) |
| *Contact* | 473 (33.6) | 415 (43.6) | 58 (12.7) |
| *Asymptomatic Screening* | 487 (34.6) | 117 (12.3) | 370 (81.3) |
| **Hospitalization**, *n (%)* | 15 (1.1) | 15 (1.6) | 0 (0.0) |

Overall, the Ag-RDT identified the presence of SARS-CoV-2 in 872 of 951 PCR-positive samples (sensitivity 91·7%; 95% CI 89·8 – 93·4 and ruled out its presence in 450 of 455 PCR-negative samples (specificity 98·9%; 95% CI 97·5 – 99·6) (Table 3). Samples with lower Ct values (i.e., a cut-off Ct <25 is associated with an increased risk of infectiousness^19–21^) showed higher sensitivity than the overall sample (Ct <25, 98.2%; Ct<30, 94.9%). Sensitivity was significantly higher among samples collected in the setting of case identification (92·6%) and contact tracing (94·2%) than asymptomatic screening (79·5%)(Table 3).

**Table 3.** Clinical performance of the Ag-RDT Panbio COVID-19 Ag Test in the overall study sample and according to RT-qPCR Ct value and disease status.

|  | SENSITIVITY |  |  |  | SPECIFICITY |  |  |  |
| --- | --- | --- | --- | --- | --- | --- | --- | --- |
|  | Ag-RDT |  |  |  | Ag-RDT |  |  |  |
|  | Detected | Not detected | PCR-positive | Sensitivity (95% CI) | Detected | Not detected | PCR-negative | Specificity (95% CI) |
| **Overall** | 872 | 79 | 951 | 91.69% (89.75-93.37) | 5 | 450 | 455 | 98.90% (97.45-99.64) |
| **Ct stratification** |  |  |  |  |  |  |  |  |
| <20 | 254 | 4 | 258 | 98.45% (96.08-99.58) | .. | .. | .. | .. |
| 20-24 | 303 | 2 | 305 | 99.34% (97.65-99.92) | .. | .. | .. | .. |
| 25-29 | 256 | 29 | 285 | 89.82% (85.71-93.08) | .. | .. | .. | .. |
| >30 | 59 | 44 | 103 | 57.28% (47.15-66.98) | .. | .. | .. | .. |
| **Disease status** |  |  |  |  |  |  |  |  |
| *Case* | 388 | 31 | 419 | 92.6% (89.66-94.92) | 0 | 27 | 27 | 100% (87.23-100) |
| <20 | 140 | 4 | 144 | 97.22% (93.04-99.24) | .. | .. | .. | .. |
| 20-24 | 129 | 1 | 130 | 99.23% (95.79-99.98) | .. | .. | .. | .. |
| 25-29 | 104 | 15 | 119 | 87.39% (80.06-92.77) | .. | .. | .. | .. |
| >30 | 15 | 11 | 26 | 57.69% (36.92-76.65) | .. | .. | .. | .. |
| *Contact* | 391 | 24 | 415 | 94.22% (91.52-96.26) | 0 | 58 | 58 | 100% (93.84-100) |
| <20 | 95 | 0 | 95 | 100% (96.19-100) | .. | .. | .. | .. |
| 20-24 | 146 | 1 | 147 | 99.32% (96.27-99.98) | .. | .. | .. | .. |
| 25-29 | 128 | 13 | 141 | 90.78% (84.75-95.00) | .. | .. | .. | .. |
| >30 | 22 | 10 | 32 | 68.75% (49.99-83.88) | .. | .. | .. | .. |
| *Asymptomatic screening* | 93 | 24 | 117 | 79.49% (71.03-86.39) | 5 | 365 | 370 | 98.65% (96.87-99.56) |
| <20 | 19 | 0 | 19 | 100% (82.35-100) | .. | .. | .. | .. |
| 20-24 | 28 | 0 | 28 | 100% (87.66-100) | .. | .. | .. | .. |
| 25-29 | 24 | 1 | 25 | 96.00% (79.65-99.90) | .. | .. | .. | .. |
| >30 | 22 | 23 | 45 | 48.89% (33.7-64.23) | .. | .. | .. | .. |

**Ag-RDT:** Ag-detecting rapid diagnostic test· **CI:** confidence interval· **Ct:** cycle threshold for RT-qPCR

However, we did not observe significant differences regarding the sensitivity estimates according to Ct value category between disease status groups (Figure 1A). In the setting of asymptomatic screening, sensitivity of samples with Ct <25 and <30 were 100% and 98.6%, respectively. All samples (except one) that tested negative for Ag-RDT in this setting had Ct values greater than 30 (Figure 1B).

**Figure 1.**
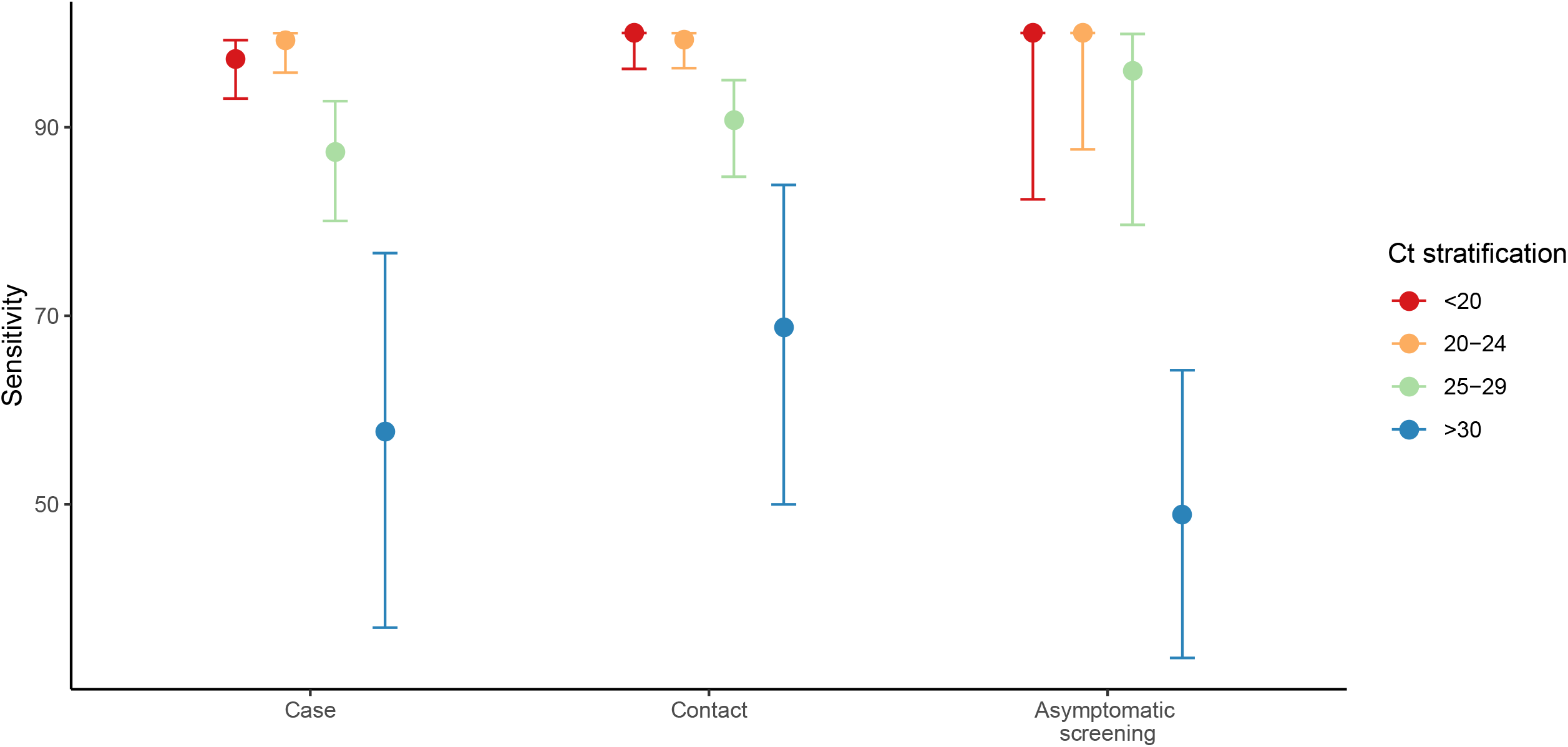
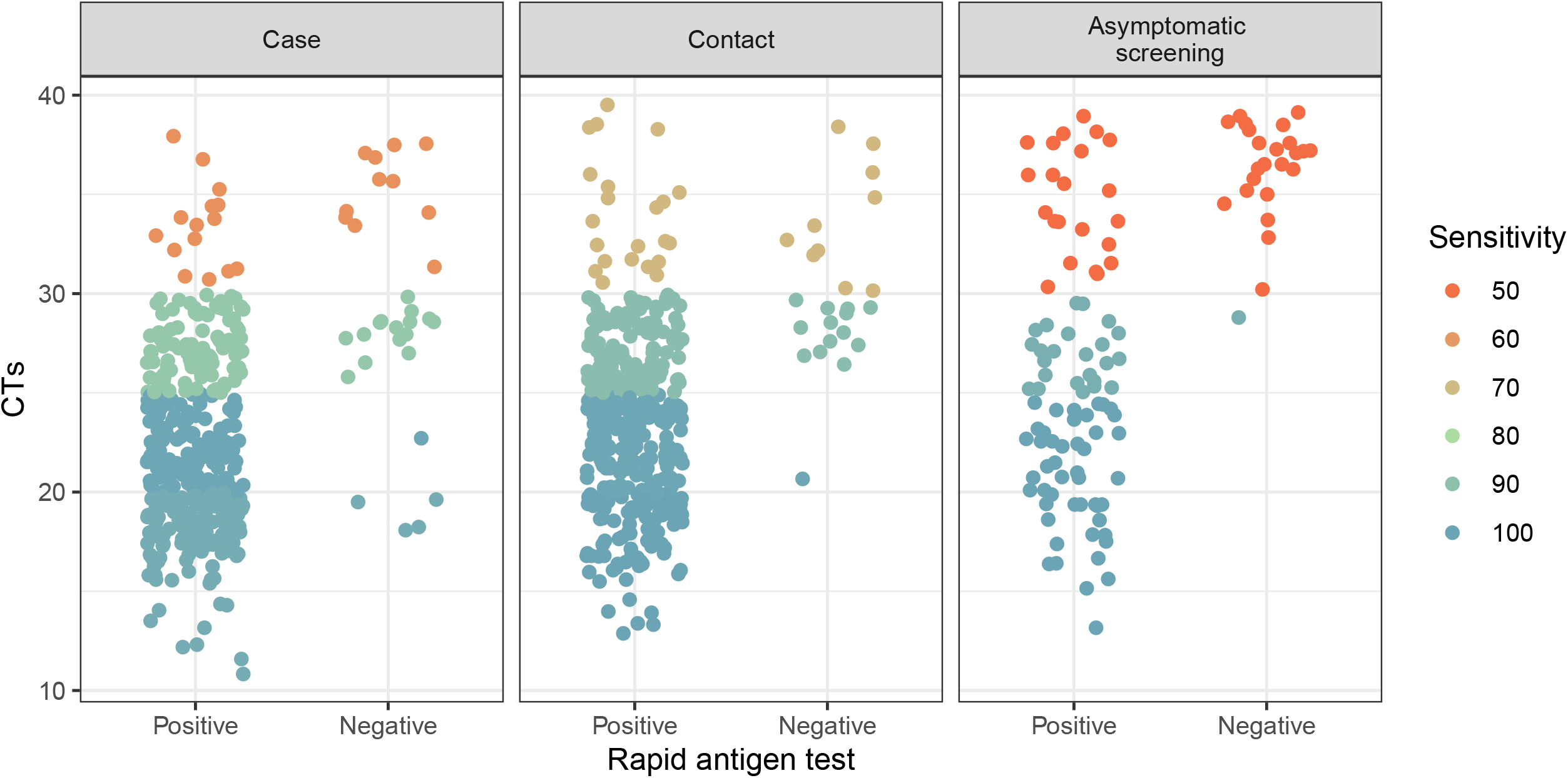
SARS-CoV-2 detection using the Ag-RDT Panbio COVID-19 Ag-Test on PCR-positive samples according to rt-qPCR Ct value. (A) Sensitivity (95CI) of the Ag-RDT according to the disease status and RT-qPCR Ct value. (B) Dot plot (individual participants) by RT-qPCR Ct value and Ag-RDT result.

Figure 2 shows the results of the modelling of the PPV and NPV based on the diagnostic performance parameters found in the overall sample. At a pre-test probability of 5%, generally assumed for asymptomatic screening in high-risk settings,^22^ the NPV was 99·6% (99·5 – 99·7) (Table 4) and increased as the pre-test probability dropped. Correspondingly, the PPV at 5% pre-test probability was 81·5% (65·0– 93·2), and decreased as pre-test probability decreased. At this pre-test probability, the estimated number of false-negative and false-positive values per thousand tests were 4 (3 – 5) and 12 (4 – 27), respectively (Figure S1, TableS2).

**Figure 2.**
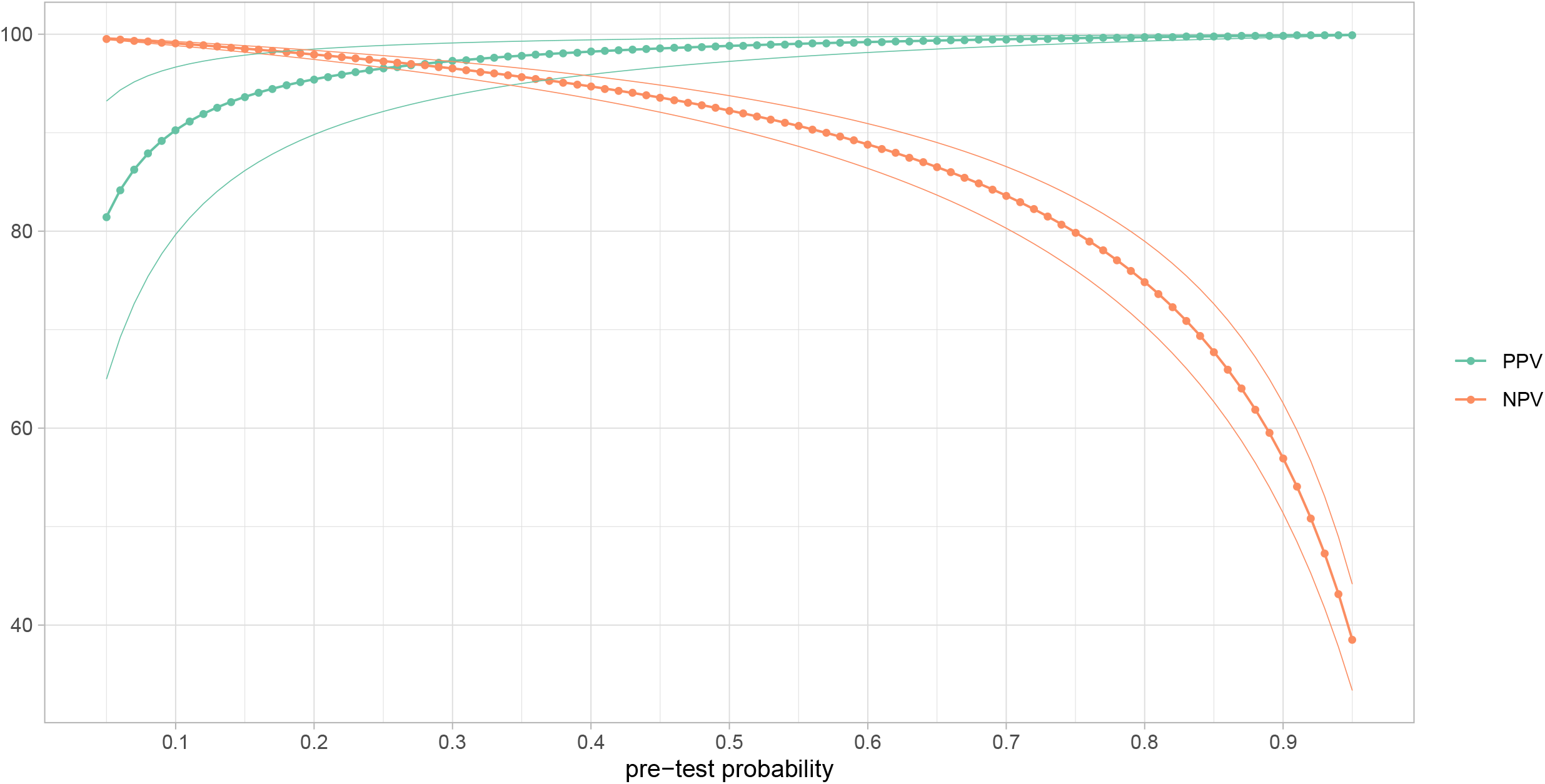
Modelling of positive predictive value (PPV) and negative predictive value (NPV) assuming different pre-test probabilities. Dots represent the PPV and NPV at sequential increment of 0·01; lines are the 95% confidence interval.

## Discussion

We showed that the point-of-care diagnostic Panbio COVID-19 Ag Test had 91·7% sensitivity and 98·9% specificity compared to standard laboratory-based RT-qPCR. Sensitivity was 99·0% for samples with Ct <25 corresponding to people at the peak of their infection, when the viral load is highest and most contagious. In nasal swab specimens from asymptomatic individuals tested in the setting of preventive screenings of the general population, sensitivity was 74·5%, although it increased to 98·9% among samples with a Ct value of <25 in this setting.

The high sensitivity of this Ag-RDT observed at higher viral loads is extremely relevant for the use of this test as a tool for epidemiological control of the SARS-CoV-2 spread because of the accumulating evidence on the high infectiousness of respiratory specimens with viral loads above 10^6^ genome copies/mL (which usually correspond to a Ct of approximately <25).^1,2,23^ Various studies have identified the Ct value of 25 as a threshold below which only a small proportion of viruses can be cultured (25% for Ct>30, 8% for Ct>35).^19^ Also, when looking at contact tracing, the secondary attack rate increases significantly for values of Ct<25,^20^ indicating notably higher infectiousness among individuals with viral loads below this threshold.

Although the Panbio COVID-19 Ag device might overlook SARS-CoV-2 infection in individuals with very low viral load (i.e., below the LoD of 6·8 × 10^5^ SARS-CoV-2 genomic copies per reaction), it maybe suitable for identifying people with high potential for infectivity. The level of viral RNA copies rises from undetectable to millions of RNA copy numbers/mL (equivalent to Ct > 40 to Ct < 25) in the order of a day and then decreases to below an infectious level by day 10 in most patients with mild infection. For some individuals, low levels of viral RNA can remain detectable by RT-qPCR for months.^24,25^ Many people whose infection are detected during community screening using high-analytic-sensitivity RT-qPCR are no longer infectious at the time of detection. The Ag-RDT reliably identifies people with high viral loads and therefore it could be useful for screening strategies to identify and isolate asymptomatic COVID-19 people while they are still infectious. Such a test could be used in focal screening to create safe environments in social activities with high-risk of transmission (e.g., visiting relatives at nursing homes, playing sports, going to a crowded place like movie theatres, music concerts, airports). It could also be used for mass screening in communities with high transmission or even for at-home frequent use.

Our study is strengthened by the large sample size and the blinding of the technicians who processed and read the results of the Ag-RDT. On the other hand, it has the limitation of not using the test under the conditions specified by the manufacturer. Our results indicate that the test can be used on frozen samples stored in transport media, thus allowing parallel sampling for Ag-RDT and PCR. However, caution should be taken when using coloured media that may affect the background of the test thin layer. In our experience, a 1:3 dilution with the Kit buffer prevented unspecific signal of yellow-coloured transport media and provided adequate results; nevertheless, we encourage validating this type of approaches before using the test. Likewise, our study was performed on stored samples rather than in a real-life setting. Owing to this last limitation, common in other assessments of the clinical performance of RDT in general,^26^ we simulated the PPV and NPV assuming a prevalence of disease based on surveillance estimates. According to our simulation, in a low prevalence setting (i.e., 5% prevalence or below), the NPV would be very high (99·6%), and screening will result in 4 (95% CI 3 – 5) false-negative results per thousand tests; the corresponding PPV would be relatively low (81·5%), stressing the need for confirmatory testing with nucleic acid amplification techniques. Irrespective of the predictive values, one must not lose sight of the relationship between the viral load and test sensitivity, a double-edged sword that better suits this test for ensuring lack of infectivity of a subject along a limited time period following test conduct.

A widely available, quick, unexpensive and accurate test could be game-changing and dramatically reduce community transmission of the virus. Newly infected people could isolate at home, severing transmission chains, and stopping the spread of the virus. We provide evidence on the high performance of the Panbio COVID-19 Ag-RDT to screen people with symptomatic and asymptomatic SARS-CoV-2 infection at high risk of virus transmission.

## Data Availability

Data Available upon request to corresponding author

## Contributors

OM, AA, BB, CGB, IB, JV designed the study. AA, BB, MU, MCM, JR, LR performed the laboratory procedures, and organized the data. DO did statistical analysis. OM wrote the first draft with revisions and input from JR, JS, CE, GF, QB, BC, JA, MVM, CGB. All authors approved the final version.

## Acknowledgements

The authors would like to thank Gerard Carot-Sans (PhD) for providing professional medical writing support during the preparation of the manuscript. We thank Andrea Tiburcio Lara and Elisabeth Bascuñana Prieto for technical support

Bárbara Baró is a Beatriu de Pinós postdoctoral fellow granted by the Government of Catalonia’s Secretariat for Universities and Research, and by Marie Sklodowska-Curie Actions COFUND Programme (BP3, 801370)

